# Detection of Mutations Associated with Variants of Concern Via High Throughput Sequencing of SARS-CoV-2 Isolated from NYC Wastewater

**DOI:** 10.1101/2021.03.21.21253978

**Authors:** Davida S. Smyth, Monica Trujillo, Kristen Cheung, Anna Gao, Irene Hoxie, Sherin Kannoly, Nanami Kubota, Michelle Markman, Kaung Myat San, Geena Sompanya, John J. Dennehy

**Affiliations:** Department of Natural Sciences and Mathematics, Eugene Lang College of Liberal Arts at The New School, New York; Department of Biology, Queensborough Community College, The City University of New York, New York; Biology Department, Queens College, The City University of New York, New York; Biology Doctoral Program, The Graduate Center, The City University of New York, New York

**Keywords:** coronavirus, environmental microbiology, Illumina sequencing, metagenomics, NGS, sewage, virus surveillance, Variants of Concern, wastewater-based epidemiology

## Abstract

Monitoring SARS-CoV-2 genetic diversity is strongly indicated because diversifying selection may lead to the emergence of novel variants resistant to naturally acquired or vaccine-induced immunity. To date, most data on SARS-CoV-2 genetic diversity has come from the sequencing of clinical samples, but such studies may suffer limitations due to costs and throughput. Wastewater-based epidemiology may provide an alternative and complementary approach for monitoring communities for novel variants. Given that SARS-CoV-2 can infect the cells of the human gut and is found in high concentrations in feces, wastewater may be a valuable source of SARS-CoV-2 RNA, which can be deep sequenced to provide information on the circulating variants in a community. Here we describe a safe, affordable protocol for the sequencing of SARS-CoV-2 RNA using high-throughput Illumina sequencing technology. Our targeted sequencing approach revealed the presence of mutations associated with several Variants of Concern at appreciable frequencies. Our work demonstrates that wastewater-based SARS-CoV-2 sequencing can inform surveillance efforts monitoring the community spread of SARS-CoV-2 Variants of Concern and detect the appearance of novel emerging variants more cheaply, safely, and efficiently than the sequencing of individual clinical samples.

**IMPORTANCE:** The SARS-CoV-2 pandemic has caused millions of deaths around the world as countries struggle to contain infections. The pandemic will not end until herd immunity is reached, that is, when most of the population has either recovered from SARS-CoV-2 infection or is vaccinated against SARS-CoV-2. However, the emergence of new SARS-CoV-2 variants of concern threatens to erase gains. Emerging new variants may re-infect persons who have recovered from COVID-19 or may evade vaccine-induced immunity. However, scaling up SARS-CoV-2 genetic sequencing to monitor Variants of Concern in communities around the world is challenging. Wastewater-based sequencing of SARS-CoV-2 RNA can be used to monitor the presence of emerging variants in large communities to enact control measures to minimize the spread of these variants. We describe here the identification of alleles associated with several variants of concern in wastewater obtained from NYC watersheds.

## INTRODUCTION

The emergence of novel SARS-CoV-2 Variants of Concern, including B.1.1.7 from the United Kingdom and B.1.351 from South Africa, has provoked intense speculation about the future of the pandemic (1-3). Early studies suggest that these new variants may be more transmissible (4-6). Even more concerning are reports of decreased antibody-mediated neutralization of these variants (7-9). Regardless of the biological attributes of these novel variants, it is clear that behavioral interventions, public health measures, vaccinations, and reduced numbers of susceptible individuals will impose strong diversifying selection on SARS-CoV-2 to enhance transmission and/or evade host immunity (10). We should anticipate that the continued evolution of SARS-CoV-2 may result in variants that evade natural or vaccine-mediated immunity. As such, intensive monitoring of SARS-CoV-2 genetic diversity and evolution is vital to rapidly identify Variants of Concern as they emerge. Currently, most SARS-CoV-2 genetic surveillance is conducted via the genome sequencing of viral RNA obtained from clinical specimens. While occurring at a much greater rate and volume than previous epidemics, the sequencing of clinical specimens is limited by cost, coverage, quality, and throughput concerns. In developed countries, these issues are not readily apparent, but sequencing efforts in underdeveloped countries has been more restricted (11). Another disadvantage of focusing on clinical strains stems from the large number of asymptomatic or mildly symptomatic infections (12). SARS-CoV-2 sequencing efforts will suffer biases if genomic information is more frequently obtained from seriously ill patients, rather than from asymptomatic patients, and those with mild symptoms who choose to follow the CDC’s advice and convalesce at home. Wastewater-based epidemiology may provide an alternative and complementary approach to provide more representative SARS-CoV-2 genetic data at lower costs and higher throughput.

Given that SARS-CoV-2 has been detected in fecal samples (13, 14), and subsequently in wastewater, wastewater is being monitored in communities around the world to determine SARS-CoV-2 prevalence in communities (15-17). Furthermore, isolation of SARS-CoV-2 RNA from wastewater coupled with high-throughput deep sequencing provides an almost unlimited source of unbiased viral sequences, which can be used to monitor frequencies of Variants of Concern in populations (18-20). We have focused on the use of targeted sequencing of the spike genomic region known to encode Variants of Concern. Our approach, while limited to a specific region of the genome, is affordable, rapid and generates sufficient coverage to quantify known variants and to identify possible emerging ones.

Our team, in conjunction with the New York City Department of Environmental Protection, has been monitoring the genetic signal of SARS-CoV-2 in the wastewater of all 14 wastewater treatment plants in NYC, an area that encompasses a population of 8,419,000 persons, since June 2020. We developed and optimized a protocol for safe, cost-effective, and repeatable quantitation of SARS-CoV-2 copy number by RT-qPCR (21). Our protocol performed strongly in a large-scale, nationwide comparative study of the reproducibility and sensitivity of 36 methods of quantifying SARS-CoV-2 in wastewater (22). Our protocol is identified as 4S.1(H) in Table 3. We further extended the utility of our protocol by deep sequencing SARS-CoV-2 RNA isolated from wastewater samples. Here we report presence of alleles associated with different Variants of Concern at appreciable frequencies. Our findings provide support for recent observations of increasing frequencies of New York Variant of Interest B.1.526 in clinical samples (23, 24), as well as the presence of Variants of Concern from the United Kingdom, California, South Africa and Brazil (25). Furthermore, our results demonstrate the utility of wastewater-based epidemiology for the timely identification of novel variants of concern arising in communities.

## RESULTS AND DISCUSSION

### Targeted sequencing is a viable approach for identifying SARS-C0V-2 mutations

We generated cDNA from NYC wastewater samples that exhibited RT-qPCR Cts values ranging from 28 to 24 Cts corresponding to 26,443 and 1,423,339 N1 copies/L, respectively. Using this cDNA as a template, we PCR amplified a region of the receptor binding domain (RBD) of the SARS-CoV-2 Spike gene, spanning amino acid residues P410 to L513, which encompasses mutations that are found in several known Variants of Concern. A total of 420 single nucleotide variants were identified in the 45 samples sequenced (Supplementary Table 1). Coverage ranged from 1,037x – 118,737x with a mean of 23,586x (Supplementary Table 1). Across all samples, we identified 75 unique mutations resulting in amino acid substitutions, 20 unique synonymous mutations, and 18 deletions resulting in a frameshift, in the 332 bp region targeted (Supplementary Table 1).

### Mutations associated with Variants of Concern are present in NYC wastewater

The five mutations found at highest frequencies, both in terms of frequency of reads within samples and found in the most samples, were L452R, E484K, N501Y, S494P, and S477N. All five mutations are associated with known Variants of Concern (Fig. 1; Supplementary Table 2). On Jan 31^st^, we sequenced samples from two wastewater treatment plants in NYC and identified reads containing mutations L452R, S477N, E484K, S494P and N501Y in both. On February 28^th^ and March 14^th^ samples from all 14 wastewater treatment plants in NYC were sequenced, revealing the presence of a high proportion of reads containing mutations L452R, S477N, E484K, S494P and N501Y (Fig. 1). Mutation L452R is unique to Pango lineage Variants of Concern B.1.427 and B.1.429, which were first observed in California (25, 26). Mutation S477N is only found in New York Variant of Interest B.1.526 (23-25, 27). Mutation E484K has been reported in Variants of Concern B.1.1.7 from the United Kingdom, P.1 and P.2 from Brazil, and B.1.351 from South Africa, and B.1.525 and B.1.526 from New York (25). Mutation S494P is only found in Variant of Concern B.1.1.7 from the United Kingdom (25). Mutation N501Y is found in Variants of Concern B.1.1.7 from the United Kingdom, P.1 from Brazil, and B.1.351 from South Africa (25).

**Figure 1.**
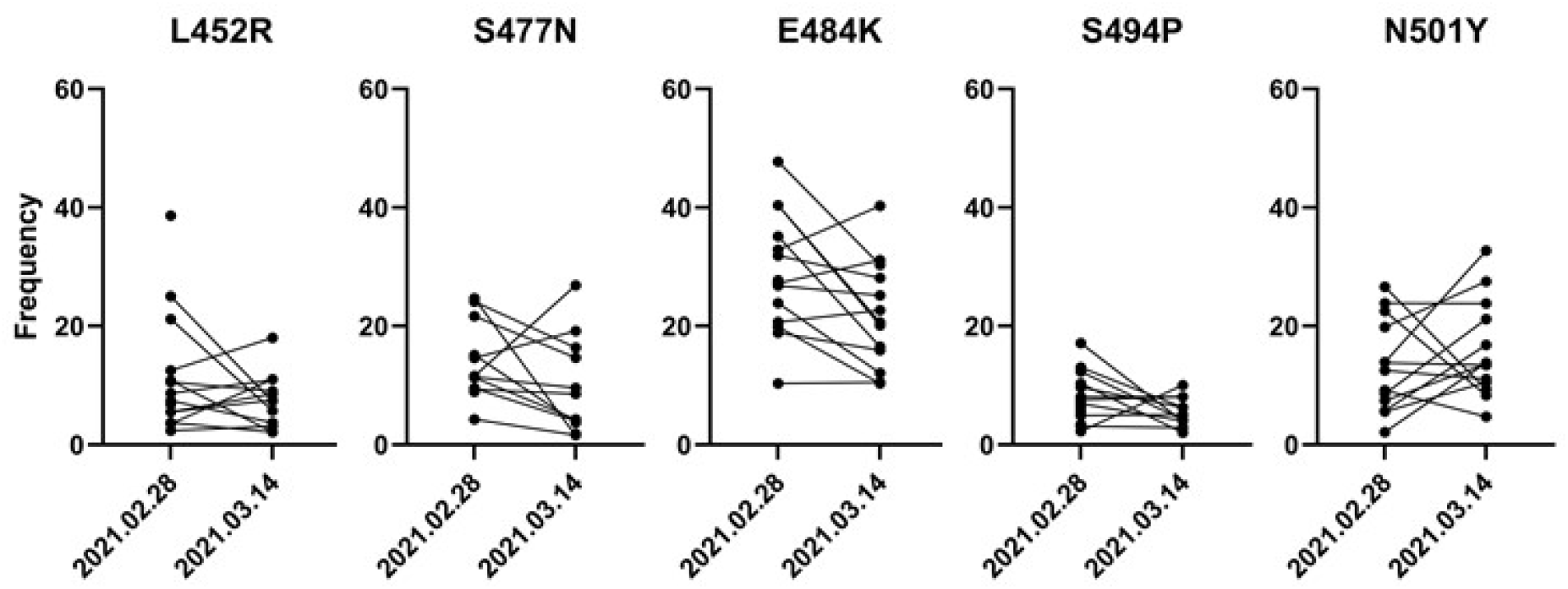
Frequencies of reads associated with five selected mutations associated with SARS-CoV-2 Variants of Concern from wastewater obtained from 14 NYC wastewater treatment plants on two separate dates.

The finding that unique mutations associated with different Variants of Concern in our pooled sequencing assay suggests the circulation of these variants in NYC. A caveat with our approach, however, is that we cannot conclusively identify the presence of a Variant of Concern since our sequencing assay targets only a region of the receptor binding domain, and some significant mutations are outside the sequenced region.

Furthermore, additional mutations occurring in the primer binding region may allow some mutations to go undetected because their DNA could not be amplified. We are expanding our targeted sequencing approach to include additional regions of interest to minimize the chance of missing important variants. Additionally, we intend to generate cDNA with random hexamers, and to incorporate a level of degeneracy in the sequencing primers to increase the breadth of our targeted sequencing.

Our most recent data from March 14^th^ suggests a slight decrease in the prevalence of the E484K variant, but we cannot draw firm conclusions due to the nature of our sequencing assay, which relies on the collective sequencing of a large pool of individuals. Nevertheless, our frequency data agrees with that recently observed in human clinical samples from NYC (23, 24, 27). We intend to supplement our targeted sequencing approach with whole genome amplicon sequencing in the future.

We believe that our approach offers a viable alternative to whole genome sequencing for the detection of known variants and can be rapidly deployed to detect additional emerging variants of concern. Importantly as a cost saving measure, labs can generate the libraries themselves and outsource the sequencing component to companies/core facilities if they lack access to a sequencer, generally with a short turnaround time.

## MATERIALS AND METHODS

### Wastewater Sample Processing and RNA Extraction

Wastewater was collected from 14 NYC wastewater treatment plants and RNA isolated according to our previously published protocol (dx.doi.org/10.17504/protocols.io.brr6m59e) (21). Control SARS-CoV-2 synthetic RNA was purchased from Twist Bioscience (#102019).

Briefly, 250 mL from 24-hr composite raw sewage samples were obtained from NYC wastewater treatment plants (WWTPs) and centrifuged at 5,000 x g for 10 min at 4°C to pellet solids. 40 mL of supernatant was passed through a 0.22 μM filter. Filtrate was stored at 4°C for 24 hrs after adding 0.9 g sodium chloride and 4.0 g PEG 8000 (Fisher) then centrifuged at 12,000 x g for 120 minutes at 4 °C to pellet precipitate. The pellet was resuspended in 1.5 mL TRIzol (Fisher), and RNA was purified according to the manufacturer’s instructions.

### Targeted PCR

Our target for sequencing was a 332 bp region of the Receptor Binding Domain (RBD) of the spike protein spanning amino acid residues P420 to L513.

Mutations in this region are of critical importance as they might help the variants evade current antibody treatments and vaccines. RNA isolated from wastewater was used to generate cDNA using ProtoScript® II Reverse Transcriptase (New England Biolabs).

The RNA was incubated with an RBD specific primer (ccagatgattttacaggctgcg) and dNTPs (0.5 mM final concentration) at 65°C for 5 minutes and placed on ice. The RT buffer, DTT (0.01 M final concentration), and the RT were added to the same tube and incubated at 42°C for 2 hours followed by 20 minutes at 65°C to inactivate the enzyme. The RBD region was amplified using Q5® High-Fidelity DNA Polymerase using the forward primer 5’ - TCGTCGGCAGCGTCAGATGTGTATAAGAGACAGccagatgattttacaggctgcg-3’ and reverse primer 5’-GTCTCGTGGGCTCGGAGATGTGTATAAGAGACAGgaaagtactactactctgtatggttgg-3’, which incorporate Illumina adaptors. PCR performed as follows: 98°C for 30 seconds, followed by 40 cycles of 98°C 5 seconds, 53°C for 15 seconds and 65°C for 1 minute and a final extension at 65°C for 1 minute.

### Targeted Sequencing

The RBD amplicons were purified using AMPure XP beads (Beckman Coulter). Index PCR was performed using the Nextera DNA CD Indexes kit (Illumina) with 2X KAPA HiFi HotStart ReadyMix (Roche), and indexed PCR products purified using AMPure beads. The indexed libraries were quantified using the Qubit 3.0 and Qubit dsDNA HS Assay Kit and diluted in 10 mM Tris-HCl to a final concentration of approximately 0.3 ng/μL (1 nM). The libraries were pooled together and diluted to a final concentration of 50 pM. Before sequencing on an Illumina iSeq100, a 10% spike-in of 50 pM PhiX control v3 (Illumina) was added to the pooled library.

### Bioinformatics

Sequencing data was uploaded to the BaseSpace Sequence Hub, and the reads demultiplexed using a FASTQ generation script. Reads were processed using the published Geneious workflows for preprocessing of NGS reads and assembly of SARS-CoV-2 amplicons (https://help.geneious.com/hc/en-us/articles/360045070991-Assembly-of-SARS-CoV-2-genomes-from-tiled-amplicon-Illumina-sequencing-using-Geneious-Prime and https://help.geneious.com/hc/en-us/articles/360044626852-Best-practice-for-preprocessing-NGS-reads-in-Geneious-Prime). Paired reads were trimmed, and the adapter sequences removed with the BBDuk plugin. Trimmed reads were merged and aligned to the SARS-CoV-2 reference genome MN908947. Variants were called using the Annotate and Predict Find Variations/SNPs in Geneious and verified by using the V-PIPE SARS-CoV-2 application (https://cbg-ethz.github.io/V-pipe/sars-cov-2/)(28).

## Supporting information

https://dennehylab.org/pw/c0nt3nt/uploads/2021/03/Supplementary-Table-1.xlsx

https://dennehylab.org/pw/c0nt3nt/uploads/2021/03/Supplementary-Table-2.docx

## Data Availability

Raw sequencing reads are available in NCBI Sequence Read Archive (SRA) under accession number PRJNA715712.

## Data Availability

Raw sequencing reads are available in NCBI’s Sequence Read Archive (SRA) under accession # PRJNA715712.

## ACKNOWLEDGEMENTS

The research described herein would not be possible if not for the assistance and support of a wide-range of organizations and individuals that came together to address the shared calamity that is the COVID-19 pandemic. We thank Gina Behnke, Esmeraldo Castro, Francoise Chauvin, Alexander Clare, Pilar Domingo-Calap, Pam Elardo, Raul Gonzalez, Crystal Hepp, Catherine Hoar, Dimitrios Katehis, William Kelly, Samantha McBride, Hope McGibbon, Hilary Millar, Samantha Patinella, Krish Ramalingam, Andrea Silverman, Jasmin Torres, Arvind Varsani, Peter Williamsen for support, advice, discussions, and feedback. This work was funded in part by the New York City Department of Environmental Protection. The Water Research Foundation, the NSF Research Coordination Network for Wastewater Surveillance for SARS-CoV-2 and Qiagen Inc. provided resources, materials and supplies, technical support, and community support.

