## Supplementary material for "Detection of Mutations Associated with Variants of Concern Via High Throughput Sequencing of SARS-CoV-2 Isolated from NYC Wastewater": https://dennehylab.org/pw/c0nt3nt/uploads/2021/03/Supplementary-Table-2.docx

| **AA Substitution** | **Frequency** | **Associated Variant of Concern** | **Comments** |
| --- | --- | --- | --- |
| L452R | 35 | B.1.427, B.1.429 |  |
| E484K | 33 | B.1.1.7, B.1.525, B.1.526 B.1.351, P.1, P.2 |  |
| N501Y | 33 | B.1.1.7, B.1.351, P.1 |  |
| S494P | 30 | B.1.1.7 |  |
| S477N | 28 | B.1.526 |  |
| Q506E | 25 | None |  |
| N427E | 14 | None |  |
| N428E | 14 | None |  |
| N440E | 13 | None |  |
| N501T | 11 | None | Frequently reported and may bind spike more strongly than wildtype. May be associated with minks. |
| T478K | 11 | B.1.1.7 |  |
| Q498H | 9 | None | Suggested to increase ACE2 binding affinity. |
| E484A | 7 | None | Possible escape mutation. |
| N501S | 6 | None | Predicted to increase infectivity. |
| Q498* | 6 | None |  |
| Q507E | 5 | None |  |
| F490Y | 4 | None |  |
| Y449R | 4 | None | Suggested to increase ACE2 binding affinity. |
| G482N | 3 | None |  |
| G485S | 3 | None |  |
| I434L | 3 | None |  |
| Q506* | 3 | None |  |
| V483F | 3 | None |  |
| A475S | 2 | None |  |
| E471G | 2 | None |  |
| F490S | 2 | None |  |
| L452Z | 2 | None |  |
| S477G | 2 | None | Suggested to increase ACE2 binding affinity. |
| T470K | 2 | None |  |
| A435S | 1 | None |  |
| A475V | 1 | None | Possible escape mutation. |
| C435T | 1 | None |  |
| E465V | 1 | None |  |
| E484Q | 1 | None |  |
| F464L | 1 | None | Possible escape mutation. |
| F486I | 1 | None | Possible escape mutation. |
| F486S | 1 | None | Possible escape mutation. |
| F490L | 1 | None | Possible escape mutation. |
| F497S | 1 | None |  |
| G446V | 1 | None | Possible escape mutation. |
| G476D | 1 | None |  |
| I468T | 1 | None | Possible increased sensitivity to mABs. |
| K444N | 1 | None | Suggested to increase infectivity. |
| K444T | 1 | None | Possible escape mutation. |
| K458E | 1 | None |  |
| K458N | 1 | None | Possible escape mutation. |
| K458R | 1 | None | Suggested to increase infectivity. |
| L441F | 1 | None |  |
| L455F | 1 | None |  |
| L455M | 1 | None |  |
| L455V | 1 | None |  |
| L461I | 1 | None |  |
| L461P | 1 | None |  |
| L462R | 1 | None |  |
| L492S | 1 | None |  |
| N440S | 1 | None |  |
| N440Y | 1 | None |  |
| N448S | 1 | None |  |
| N450D | 1 | None | Possible escape mutation. |
| N460L | 1 | None |  |
| N487D | 1 | None |  |
| P499H | 1 | None |  |
| Q493L | 1 | None |  |
| S438T | 1 | None |  |
| S443P | 1 | None |  |
| S459T | 1 | None |  |
| S469L | 1 | None |  |
| S469P | 1 | None |  |
| S494L | 1 | None |  |
| V433A | 1 | None |  |
| V433F | 1 | None |  |
| V445A | 1 | None |  |
| W436R | 1 | None |  |
| Y473H | 1 | None |  |
| Y495H | 1 | None |  |
